# Progression of chronic kidney disease among black patients attending a tertiary hospital in Johannesburg, South Africa

**DOI:** 10.1101/2022.10.06.22280771

**Authors:** Alfred Meremo, Graham Paget, Raquel Duarte, Deogratius Bintabara, Saraladevi Naicker

## Abstract

**Background:** Chronic kidney disease (CKD) is a major public health issue worldwide and is an important contributor to the overall non-communicable disease burden. Chronic kidney disease is usually asymptomatic, and insidiously and silently progresses to advanced stages in resource limited settings.

**Methodology:** A prospective longitudinal study was carried out on black patients with CKD attending the kidney outpatient clinic at Charlotte Maxeke Johannesburg Academic Hospital (CMJAH) in South Africa, between September 2019 to March 2022. Demographic and clinical data were extracted from the ongoing continuous clinic records, as well as measurements of vital signs and interviews at baseline and at follow up. Patients provided urine and blood samples for laboratory investigations as standard of care at study entry (0) and at 24 months, and were followed up prospectively for two (2) years. Data were descriptively and inferentially entered into REDcap and analysed using STATA version 17, and multivariable logistic regression analysis was used to identify predictors of CKD progression.

**Results:** A total of 312 patients were enrolled into the study, 297 (95.2%) patients completed the study, 10 (3.2%) patients were lost to follow and 5 (1.6%) patients died during the study period. The prevalence of CKD progression was 49.5%, while that of CKD remission was 33% and CKD regression was 17.5%. For patients with CKD progression the median age at baseline was 58 (46 - 67) years, the median eGFR was 37 (32 -51) mL/min/1.73 m^2^, median urine protein creatinine ratio (uPCR) was 0.038 (0.016 -0.82) g/mmol and the median haemoglobin (Hb) was 13.1 (11.7 – 14.4) g/dl; 95.2% had hypertension, 40.1% patients had diabetes mellitus and 39.5% had both hypertension and diabetes mellitus. Almost half (48.3%) of patients with CKD progression had severely increased proteinuria and 45.6% had anaemia. Variables associated with higher odds for CKD progression after multivariable logistic regression analysis were severely increased proteinuria (OR 32.3, 95 % CI 2.8 - 368.6, P = 0.005), moderately increased proteinuria (OR 23.3, 95% CI 2.6 - 230.1, P= 0.007), hypocalcaemia (OR 3.8, 95 % CI 1.0 - 14.8, P = 0.047), hyponatraemia (OR 4.5, 95% CI 0.8 - 23.6, P= 0.042), anaemia (OR 2.1, 95% CI 1.0 - 4.3, P= 0.048), diabetes mellitus (OR 1.8, 95 % CI 0.9 - 3.6, P = 0.047), elevated HbA1c (OR 1.8, 95 % CI 1.2 - 2.8, P = 0.007) and current smoking (OR 2.8, 95 % CI 0.9 - 8.6, P = 0.049).

**Conclusion:** Our study identified a higher prevalence of progression of CKD in a prospective longitudinal study of black patients with CKD. Progression of CKD was associated with proteinuria, diabetes mellitus, elevated HbA1c, anaemia, hypocalcaemia, hyponatraemia and current smoking. This is a call for nephrologists and clinicians to be vigilant in identifying CKD patients at risk of CKD progression at early stages as this would allow risk stratification to improve kidney disease outcomes.

## Introduction

The global prevalence of chronic kidney disease (CKD) has been progressively increasing and is a significant risk factor for cardiovascular disease morbidity and mortality (1, 2). The burden of CKD is more prominent in low- and lower-middle-income countries (LLMICs) than in high-income countries (HICs) with a higher prevalence among those disadvantaged and indigenous communities who have less/ limited access to health care (1, 3). The global increase in CKD is driven by the increasing prevalence of diabetes mellitus, hypertension and obesity (4, 5). Studies have shown that African-Americans have a 2-to 4-fold higher risk for end-stage kidney disease (ESKD) than their white counterparts requiring renal replacement therapy (6, 7). Individuals of black ethnicity due to their genes, including the presence of *APOL*1 high-risk genotypes, are at higher risk of increased serum creatinine levels, lower eGFR, more rapid CKD progression and death due to social, economic and medical inequities (8, 9). Studies have reported the prevalence of CKD to be 15.8% in Africa, with major cost implications to overburdened healthcare systems (10, 11). In developing countries, patients with ESKD are increasing in numbers and most die due to poor access to kidney replacement therapies, calling for cost effective preventive strategies to reduce CKD progression (4, 12). Chronic kidney disease in developing countries also tends to be present in younger people who are unaware of their underlying kidney disease, and very few receive optimal care because of resource constraints (13, 14). Regardless of the underlying cause of CKD, most kidney diseases progress to ESKD due to irreversible nephron loss leading to premature death (15, 16). Slowing CKD progression, promoting remission, or even regression of CKD, have been based on the control of blood pressure, control of blood sugar, reduction of proteinuria and management of other known risk factors for CKD progression (17, 18). When CKD progressors are identified reliably and earlier in stages 1 to 3, disease progression can be altered with reduced complications (19, 20). There is a need to identify those patients with CKD and those who are likely to progress more rapidly, to reduce the number of patients progressing to end-stage kidney disease (ESKD). The aim of this study was to study the prevalence and predictors of CKD progression in a cohort of black patients with satisfactory blood pressure and glycaemic control attending a tertiary hospital in Johannesburg, South Africa.

## Methods

### Study design, population and settings

This was a prospective longitudinal study to evaluate the prevalence and predictors of CKD progression among black patients attending the kidney outpatient clinic at the Charlotte Maxeke Johannesburg Academic Hospital (CMJAH) between September 2019 to March 2022. The CMJAH is a public accredited central hospital with 1088 beds serving patients from across the Gauteng province and nearby provinces in South Africa. CMJAH is also one of the main teaching hospitals for the Faculty of Health Sciences of the University of the Witwatersrand. Johannesburg is the largest city in South Africa and among the largest 50 urban agglomerations in the world. Johannesburg had an estimated population of around 5.9 million in 2021, the most common racial groups include black African (76.4%), White (12.3%), mixed ethnicity (5.6%) and Indian/Asian (4.9%). Inclusion criteria for the study were patients who were >18 years of age, with CKD stages 1 – 4, who had controlled hypertension (blood pressure < 140/90 mm Hg) and diabetes mellitus (HbA1C < 7%), attending the kidney outpatient clinic for at least 6 months and were able to provide informed consent. Patients who had active infections, active malignancies, autoimmune diseases and who were not black were excluded. Black patients have significantly higher rates of more rapid CKD progression and ESKD than other races (21, 22), hence they were the focus of this study.

### Data collection and laboratory procedures

Enrollment occurred between September 2019 to March 2020; consecutive sampling was used to meet the required number of study patients and follow up was conducted between September 2021 to March 2022. Demographic and clinical data including age, gender, weight, height, glycaemic status, history of smoking, aetiology of CKD and medications were extracted from the ongoing continuous clinic records. Face to face interviews at the time of enrolment were recorded in a questionnaire and vital signs measurements were conducted at first enrolment and at follow up. Patients provided blood and urine samples at study entry (0) and at 24 months; patients agreed to have their routine medical records accessed prospectively for two (2) years. Systolic and diastolic blood pressure was measured 3 times, and the average of the second and third measurements was used. Body mass index (BMI) was calculated using the National Health Services (NHS-UK) BMI calculator (23). Measurements of urinary protein creatinine ratio (uPCR), serum creatinine, electrolytes, HbA1C, white cell count, haemoglobin level, platelets, calcium, phosphate, transferrin and HDL cholesterol were done as standard of care at the time of recruitment and at follow up during a clinic visit. Anaemia was defined as a Hb concentration of < 13.0g/dl in males and < 12.0g/dl in females (24). Serum creatinine was measured using the isotope dilution mass spectrometry (IDMS) traceable enzymatic assay and estimated glomerular filtration rate (eGFR) was calculated using the Chronic Kidney Disease Epidemiology Collaboration (CKD-EPI) equation without using the African American correction factor (25). CKD progression was assessed by a (i) change to a more advanced stage of CKD (ii) greater than 30% reduction of eGFR in two years, whereby percent change in eGFR was calculated as last eGFR – first eGFR/ first eGFR x 100 (26) (iii) sustained decline in eGFR of > 4 ml/min/1.73 m^2^/year or more (27, 28). CKD regression was defined by a sustained increase in eGFR of > 4 ml/min/1.73 m^2^/year or an improvement in CKD stage at 24 months from baseline (18, 27). CKD remission was defined by a stable or change in eGFR of < 4 ml/min/1.73 m^2^/year from baseline and at 24 months of follow up (18, 28).

### Data management and analysis

Study data were collected and entered into *REDCap* (Research Electronic Data Capture) tools (29, 30) hosted at the University of the Witwatersrand and analyzed using STATA version 17 (College Station, Texas, USA). Descriptive statistics were used to summarize demographic and clinical data; continuous variables have been reported as medians with interquartile ranges and Wilcoxon rank-sum test was used for the non-normally distributed variables. Discrete variables have been reported as frequencies and proportions, Pearson’s chi-square test were used to test for association between variables and CKD progression. Odd ratios were used to estimate the strengths of association between variables and CKD progression; univariate and multivariate logistic regression models were used to estimate the association of variables and CKD progression. Variables with p-value less than 0.2 on univariate logistic regression models were then fitted into the multivariate logistic regression models with the addition of age and sex as adjusting variables; variables with a p-value of less than 0.05 were considered to have significant strength of association.

### Ethical issues

Ethical approval was obtained from the Human Research Ethics Committee of the University of Witwatersrand, Johannesburg (ethics clearance certificate No. M190553). Written informed consent was obtained from each of the participants before enrolment and embarking on data collection.

## Results

### Demographic and clinical characteristics of the study population

A total of 476 black patients with CKD stages 1-4 attended the CMJAH kidney outpatient clinic during the enrolment period, 164 patients were excluded from the study including 110 patients who had uncontrolled hypertension, 35 patients who had uncontrolled diabetes mellitus, 11 patients who had autoimmune diseases, 6 patients had active infections and 2 patients had active malignancies. Of the 312 patients who were enrolled into the study, 10 (3.2%) patients were lost to follow and 5 (1.6%) patients died, 297 (95.2%) patients completed the study of whom 156 (52.5 %) were male and 154 (51.8 %) were married. Progression of CKD occurred in 147 (49.5%) patients over 2 years of follow up. The median age was 58 (46 - 67) years for CKD progressors and 57 (45 - 66) years for those without CKD progression. Significant variables were the median eGFR at baseline, 37 (32 -51) mL/min/1.73 m^2^ for CKD progressors and 44 (34 – 61) mL/min/1.73 m^2^ for those without CKD progression (p=0.003) ; the median urine protein creatinine ratio (uPCR) was 0.038 (0.016 -0.82) g/mmol for CKD progressors and 0.016 (0.008 – 0.032) g/mmol for those without CKD progression (p=0.001); median haemoglobin of 13.1 (11.7 – 14.4) g/dl for CKD progressors and 13.7 (12.2 – 15.3) g/dl for those without CKD progression, p=0.023 (Table 1).

**Table 1:**
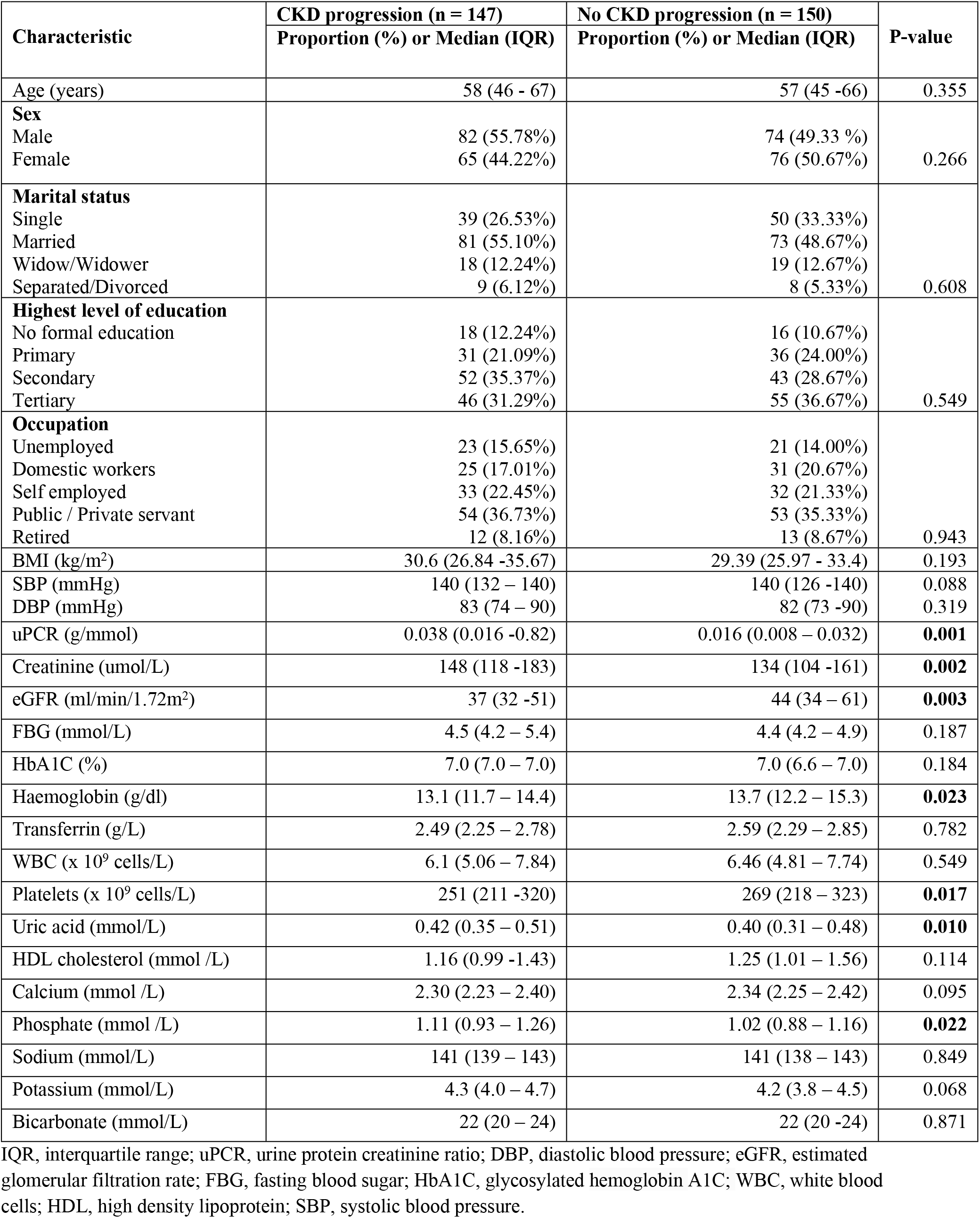
Demographic and clinical characteristics of CKD progressors and non-progressors.

| Characteristic | CKD progression (n = 147) | No CKD progression (n = 150) | P-value |
| --- | --- | --- | --- |
|  | Proportion (%) or Median (IQR) | Proportion (%) or Median (IQR) |  |
| Age (years) | 58 (46 - 67) | 57 (45 -66) | 0.355 |
| **Sex** |  |  |  |
| Male | 82 (55.78%) | 74 (49.33 %) | 0.266 |
| Female | 65 (44.22%) | 76 (50.67%) |  |
| **Marital status** |  |  |  |
| Single | 39 (26.53%) | 50 (33.33%) | 0.608 |
| Married | 81 (55.10%) | 73 (48.67%) |  |
| Widow/Widower | 18 (12.24%) | 19 (12.67%) |  |
| Separated/Divorced | 9 (6.12%) | 8 (5.33%) |  |
| **Highest level of education** |  |  |  |
| No formal education | 18 (12.24%) | 16 (10.67%) | 0.549 |
| Primary | 31 (21.09%) | 36 (24.00%) |  |
| Secondary | 52 (35.37%) | 43 (28.67%) |  |
| Tertiary | 46 (31.29%) | 55 (36.67%) |  |
| **Occupation** |  |  |  |
| Unemployed | 23 (15.65%) | 21 (14.00%) | 0.943 |
| Domestic workers | 25 (17.01%) | 31 (20.67%) |  |
| Self employed | 33 (22.45%) | 32 (21.33%) |  |
| Public / Private servant | 54 (36.73%) | 53 (35.33%) |  |
| Retired | 12 (8.16%) | 13 (8.67%) |  |
| BMI (kg/m<sup>2</sup>) | 30.6 (26.84 -35.67) | 29.39 (25.97 - 33.4) | 0.193 |
| SBP (mmHg) | 140 (132 – 140) | 140 (126 -140) | 0.088 |
| DBP (mmHg) | 83 (74 – 90) | 82 (73 -90) | 0.319 |
| uPCR (g/mmol) | 0.038 (0.016 -0.82) | 0.016 (0.008 – 0.032) | **0.001** |
| Creatinine (umol/L) | 148 (118 -183) | 134 (104 -161) | **0.002** |
| eGFR (ml/min/1.72m<sup>2</sup>) | 37 (32 -51) | 44 (34 – 61) | **0.003** |
| FBG (mmol/L) | 4.5 (4.2 – 5.4) | 4.4 (4.2 – 4.9) | 0.187 |
| HbA1C (%) | 7.0 (7.0 – 7.0) | 7.0 (6.6 – 7.0) | 0.184 |
| Haemoglobin (g/dl) | 13.1 (11.7 – 14.4) | 13.7 (12.2 – 15.3) | **0.023** |
| Transferrin (g/L) | 2.49 (2.25 – 2.78) | 2.59 (2.29 – 2.85) | 0.782 |
| WBC (x 10<sup>9</sup> cells/L) | 6.1 (5.06 – 7.84) | 6.46 (4.81 – 7.74) | 0.549 |
| Platelets (x 10<sup>9</sup> cells/L) | 251 (211 -320) | 269 (218 – 323) | **0.017** |
| Uric acid (mmol/L) | 0.42 (0.35 – 0.51) | 0.40 (0.31 – 0.48) | **0.010** |
| HDL cholesterol (mmol /L) | 1.16 (0.99 -1.43) | 1.25 (1.01 – 1.56) | 0.114 |
| Calcium (mmol /L) | 2.30 (2.23 – 2.40) | 2.34 (2.25 – 2.42) | 0.095 |
| Phosphate (mmol /L) | 1.11 (0.93 – 1.26) | 1.02 (0.88 – 1.16) | **0.022** |
| Sodium (mmol/L) | 141 (139 – 143) | 141 (138 – 143) | 0.849 |
| Potassium (mmol/L) | 4.3 (4.0 – 4.7) | 4.2 (3.8 – 4.5) | 0.068 |
| Bicarbonate (mmol/L) | 22 (20 – 24) | 22 (20 -24) | 0.871 |

IQR, interquartile range; uPCR, urine protein creatinine ratio; DBP, diastolic blood pressure; eGFR, estimated glomerular filtration rate; FBG, fasting blood sugar; HbA1C, glycosylated hemoglobin A1C; WBC, white blood cells; HDL, high density lipoprotein; SBP, systolic blood pressure.

### Prevalence of CKD progression, CKD remission and CKD regression

During the 2 years of follow up, a total of 147 (49.5%) patients had progression of CKD whereby 35.0% changed to a more advanced stage of CKD, 19.9% had greater than 30% reduction of eGFR in two years and 48.5 % had a sustained decline in eGFR of > 4 ml/min/1.73 m^2^/year. A total of 98 (33%) patients were observed to have remission of CKD, while 52 (17.5%) patients had CKD regression (Figures 1a & 1b).

**Figure 1a:**
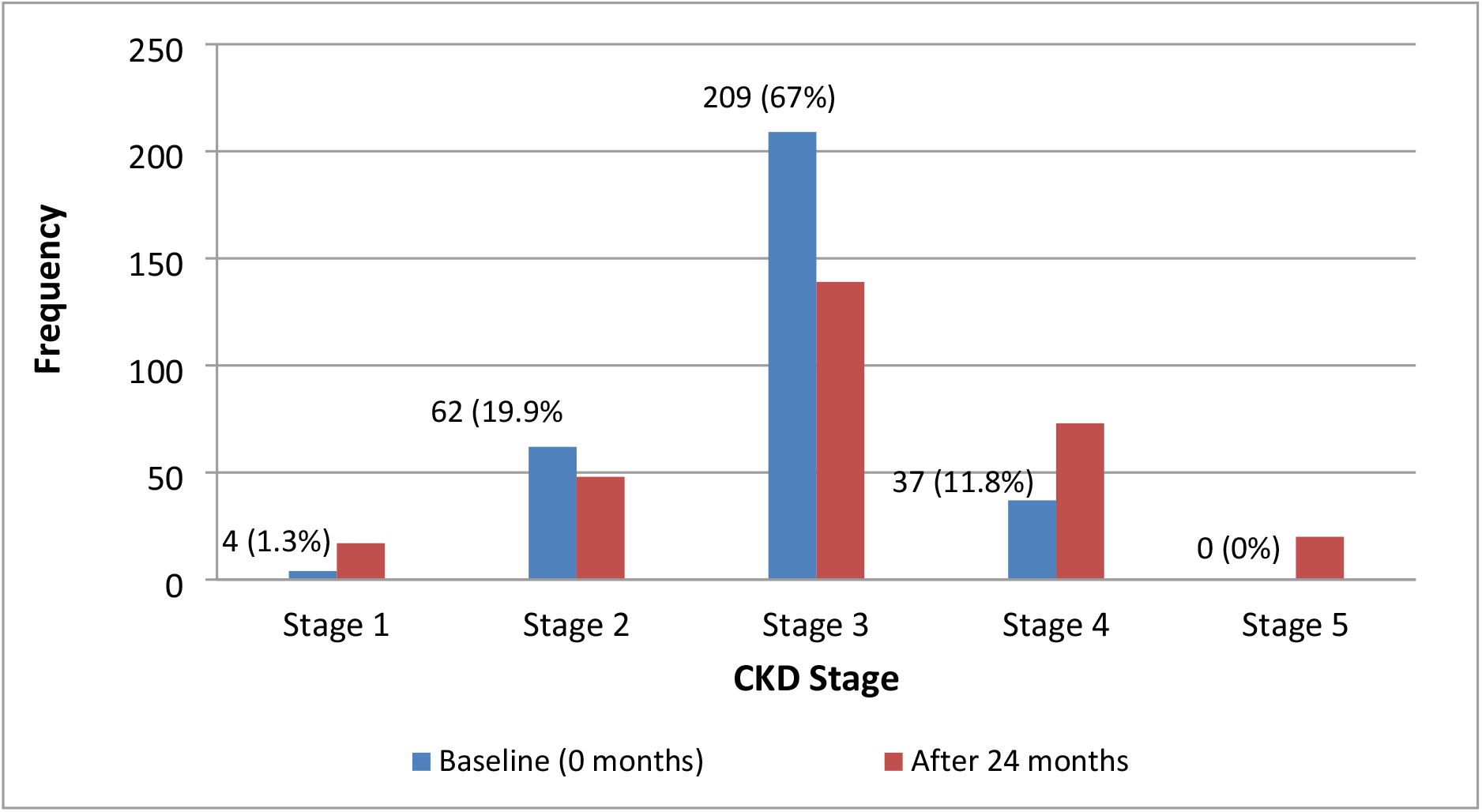
CKD stages at baseline and at 24 months follow up

**Figure 1b:**
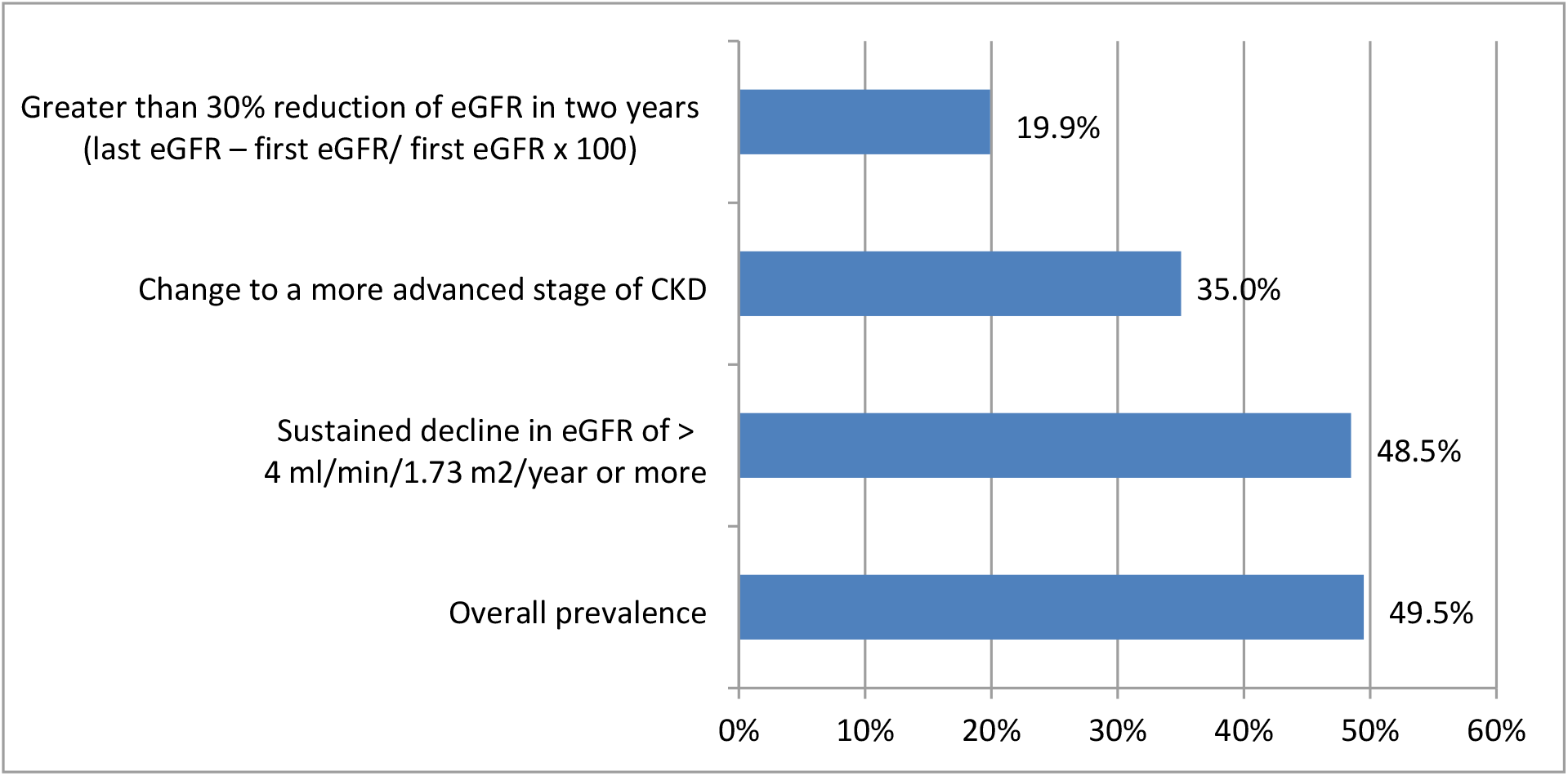
Estimation of CKD progression using different eGFR criteria

### Clinical profile of black patients who had CKD progression

Of the 147 black patients who had CKD progression, the majority (95.2%) of patients had hypertension, 59 (40.1%) patients had diabetes mellitus and 58 (39.5%) had both hypertension and diabetes mellitus. About half (48.3%) of the CKD progressors presented with severely increased proteinuria as compared to 30 (20.0%) of those without CKD progression. Anaemia was noted in 67 (45.6%) patients with CKD progression and in 40 (26.7 %) patients in those without CKD progression. Low serum calcium levels were noted in 26 (17.7%) of those with CKD progression and in 10 (6.7%) patients of those without CKD progression, while high serum phosphate levels were noted in 18 (12.3%) patients in those with CKD progression and in 6 (4.0%) patients of those without CKD progression. Low serum sodium levels were noted in 12 (8.2%) of those with CKD progression and in 3 (2.0%) of patients without CKD progression, while high serum potassium levels were noted in 21 (14.3%) patients who had CKD progression and in 7 (4.7%) of patients without CKD progression. Hyperuricemia was present in 50 (34.0%) patients with CKD progression as compared to 24 (16.0%) patients without CKD progression. Majority (76.9%) patients with CKD progression were using more than 4 anti-hypertensives for their blood pressure control as compared to 57.3 % in those patients without CKD progression. For patients with CKD progression, the majority (85.7%) of patients were on calcium channel blockers, 62.6% were on diuretics, 57.1% were on statins, 55.1% were on beta blockers, 31.3% were on insulin, 29.3% and were on aspirin. A total of 35 (23.8 %) CKD progressors were on ACEs/ARBs compared with 25 (16.7 %) non-progressors; there was no significant difference in CKD progression associated with the use of ACEIs/ARBs (Table 2).

**Table 2:** Clinical and laboratory parameters of CKD patients.

| Parameter | CKD progression (n = 147) Proportion (%) | No CKD progression (n = 150) Proportion (%) | P-value |
| --- | --- | --- | --- |
| **eGFR (ml/min/1.72m<sup>2</sup>)** |  |  |  |
| Stage 1 (> 90) | 0 (0.0 %) | 17 (11.3 %) | **0.001** |
| Stage 2 (60 – 89) | 13 (8.84 %) | 35 (23.33 %) |  |
| Stage 3 (30 -59) | 50 (34.01 %) | 89 (59.33 %) |  |
| Stage 4 (16- 29) | 64 (43.54 %) | 9 (6.0 %) |  |
| Stage 5 (< 15) | 20 (13.61%) | 0 (0.0 %) |  |
| **Diagnoses encountered** |  |  |  |
| Hypertension | 140 (95.24 %) | 137 (91.33 %) | 0.179 |
| Diabetes mellitus | 59 (40.14 %) | 39 (26.00 %) | **0.010** |
| Hypertension & Diabetes mellitus | 58 (39.46%) | 38 (25.3%) | **0.009** |
| Adult polycystic kidney disease | 7 (4.76 %) | 4 (2.67 %) | 0.339 |
| Reflux nephropathy | 2 (1.36 %) | 3 (2.00 %) | 0.668 |
| Obstructive uropathy | 1 (0.68 %) | 0 (0.0 %) | 0.312 |
| Unknown | 2 (1.36 %) | 8 (5.33 %) | 0.058 |
| **Current smoking** |  |  |  |
| Yes | 14 (9.52%) | 8 (5.33 %) | 0.168 |
| No | 133 (90.48%) | 142 (96.67%) |  |
| **Current Alcohol** |  |  |  |
| Yes | 14 (9.52%) | 18 (12.00%) | 0.493 |
| No | 133 (90.48%) | 132 (88.00%) |  |
| **Cardiovascular Diseases** |  |  |  |
| None | 99 (67.35%) | 121 (80.67 %) | **0.030** |
| Angina | 28 (19.05%) | 22 (14.67 %) |  |
| Myocardial infarction | 9 (6.12%) | 2 (1.33 %) |  |
| Heart failure | 5 (3.40%) | 3 (2.00 %) |  |
| Stroke | 6 (4.08%) | 2 (1.33 %) |  |
| **Medications** |  |  |  |
| None | 4 (2.72 %) | 5 (3.33 %) | 0.758 |
| Diuretics | 92 (62.59 %) | 61 (40.67 %) | **0.001** |
| ACEIs / ARBs | 35 (23.81 %) | 25 (16.67 %) | 0.125 |
| Aldactone | 7 (4.76 %) | 4 (2.67 %) | 0.339 |
| CCBs | 126 (85.71 %) | 112 (74.67 %) | **0.017** |
| Statins | 84 (57.14 %) | 67 (44.67 %) | **0.032** |
| Oral hypoglycemics | 15 (10.2 %) | 21 (14.00 %) | 0.316 |
| Insulin | 46 (31.29 %) | 22 (14.67 %) | **0.001** |
| Allopurinol | 16 (10.88 %) | 18 (12.00 %) | 0.763 |
| Junior ASA | 43 (29.25 %) | 26 (17.33 %) | **0.015** |
| Beta blockers | 81 (55.10 %) | 61 (40.67 %) | **0.013** |
| Aldomet | 36 (24.49 %) | 33 (22.0 %) | 0.611 |
| Hydralazine | 11 (7.48 %) | 8 (5.33 %) | 0.449 |
| Nitrates (ISMN/ISDN) | 4 (2.72%) | 3 (2.00%) | 0.682 |
| Doxazosin | 68 (46.26 %) | 66 (44.00 %) | 0.696 |
| Others | 47 (31.97 %) | 52 (34.67 %) | 0.622 |
| **No. of antihypertensives per patient** |  |  |  |
| 0 | 4 (2.72 %) | 6 (4.00 %) | **0.002** |
| 1 - 3 | 30 (20.40 %) | 58 (38.67 %) |  |
| ≥ 4 | 113 (76.88 %) | 86 (57.33 %) |  |
| **uPCR (g/mmol)** |  |  |  |
| Normal to mildly increased (<0.015) | 23 (15.65 %) | 59 (39.33 %) | **0.001** |
| Moderately increased (0.015- 0.05) | 48 (32.65 %) | 59 (39.33 %) |  |
| Severely increased (> 0.050) | 71 (48.30 %) | 30 (20.00 %) |  |
| Nephrotic syndrome (> 0.350) | 5 (3.40%) | 2 (1.33%) |  |
| **WBC (x 10<sup>9</sup> cells/l)** |  |  |  |
| Low (> 4) | 5 (3.40 %) | 12 (8.00 %) | 0.096 |
| Normal (4-11) | 139 (94.56 %) | 131 (87.33 %) |  |
| High (>11) | 3 (2.04 %) | 7 (4.67 %) |  |
| **Haemoglobin (g/dl)** |  |  |  |
| Normal (> 12.0 or 13.0) | 80 (54.42 %) | 110 (73.33 %) | **0.001** |
| Anemia (<12.0 or 13.0) | 67 (45.58 %) | 40 (26.67 %) |  |
| **Transferrin g/dl)** |  |  |  |
| Low (< 2.0) | 26 (17.69 %) | 13 (8.67 %) | 0.061 |
| Normal (2.0-3.60) | 117 (79.59 %) | 134 (89.33 %) |  |
| High (>3.60) | 4 (2.72 %) | 3 (2.00 %) |  |
| **Platelets (x 10<sup>9</sup> cells/l)** |  |  |  |
| Low (> 150) | 2 (1.36 %) | 3 (2.00 %) | 0.643 |
| Normal (150 - 450) | 142 (96.60 %) | 141 (94.00 %) |  |
| High (>450) | 3 (2.04 %) | 6 (4.00 %) |  |
| **Calcium (mmol /l)** |  |  |  |
| Low (< 2.15) | 26 (17.69 %) | 10 (6.67 %) | **0.009** |
| Normal (2.15 -2.45) | 107 (72.79 %) | 118 (78.67 %) |  |
| High (>2.45) | 14 (9.52 %) | 22 (14.67 %) |  |
| **Phosphate (mmol /l)** |  |  |  |
| Low (< 0.80) | 7 (4.76 %) | 16 (10.67 %) |  |
| Normal (0.80 -1.55) | 122 (82.99 %) | 128 (85.33 %) |  |
| High (>1.55) | 18 (12.25 %) | 6 (4.00 %) | **0.019** |
| **Uric acid (mmol/l)** |  |  |  |
| Normal (0.16/0.21 – 0.36/0.43) | 97 (65.99 %) | 126 (84.00 %) | **0.001** |
| High (> 0.36 or 0.43) | 50 (34.01 %) | 24 (16.00 %) |  |
| **HDL cholesterol (mmol /l)** |  |  |  |
| Low (< 1.04) | 43 (29.25 %) | 35 (23.33 %) | 0.343 |
| Normal (1.04 -1.17) | 28 (19.05 %) | 25 (16.67 %) |  |
| High (>1.17) | 76 (51.70 %) | 90 (60.00 %) |  |
| **Sodium (mmol/l)** |  |  |  |
| Low (< 136) | 12 (8.16 %) | 3 (2.00 %) | **0.034** |
| Normal (136 -145) | 130 (88.44 %) | 138 (92.00 %) |  |
| High (>145) | 5 (3.40 %) | 9 (6.00 %) |  |
| **Potassium (mmol/l)** |  |  |  |
| Low (< 3.5) | 10 (6.80 %) | 11 (7.33 %) | **0.018** |
| Normal (3.5 – 5.1) | 116 (78.91 %) | 132 (88.0 %) |  |
| High (> 5.1) | 21 (14.29 %) | 7 (4.67 %) |  |
| **Bicarbonate (mmol/l)** |  |  |  |
| Low (< 23) | 97 (65.99 %) | 85 (56.67 %) | 0.130 |
| Normal (23 – 29) | 46 (31.29 %) | 63 (42.00 %) |  |
| High (> 29) | 4 (2.72 %) | 2 (1.33%) |  |

uPCR, urine protein creatinine ratio; eGFR, estimated glomerular filtration rate; CCBs, calcium channel blockers; ACEIs, angiotensin converting enzyme inhibitors; ARBs, Aldosterone receptors blockers.

### Predictors of CKD progression

We divided the patients into two subgroups according to their CKD progression status after 2 years of follow up: CKD progression vs. no CKD progression. A total of 30 potential variables were identified after performing univariate logistic regression analyses. Backward elimination reduced this to 21 parameters; the factors associated with CKD progression after adjusting for age and sex on multivariate logistic regression analysis included diabetes mellitus (OR 1.8, 95 % CI 0.9 - 3.6, P = 0.047), increasing HbA1C (OR 1.8, 95 % CI 1.2 - 2.8, P = 0.007), severely increased proteinuria (OR 32.3, 95 % CI 2.8 - 368.6, P = 0.005), moderately increased proteinuria (OR 23.3, 95% CI 2.6 - 230.1, P=0.007), hypocalcaemia (OR 3.8, 95 % CI 1.0 - 14.8, P = 0.047), anaemia (OR 2.1, 95% CI 1.0 - 4.3, P= 0.048), hyponatraemia (OR 4.5, 95% CI 0.8 - 23.6, P= 0.042) and current smoking (OR 2.8, 95 % CI 0.9 - 8.6, P = 0.049 (Tables 3).

**Table 3:**
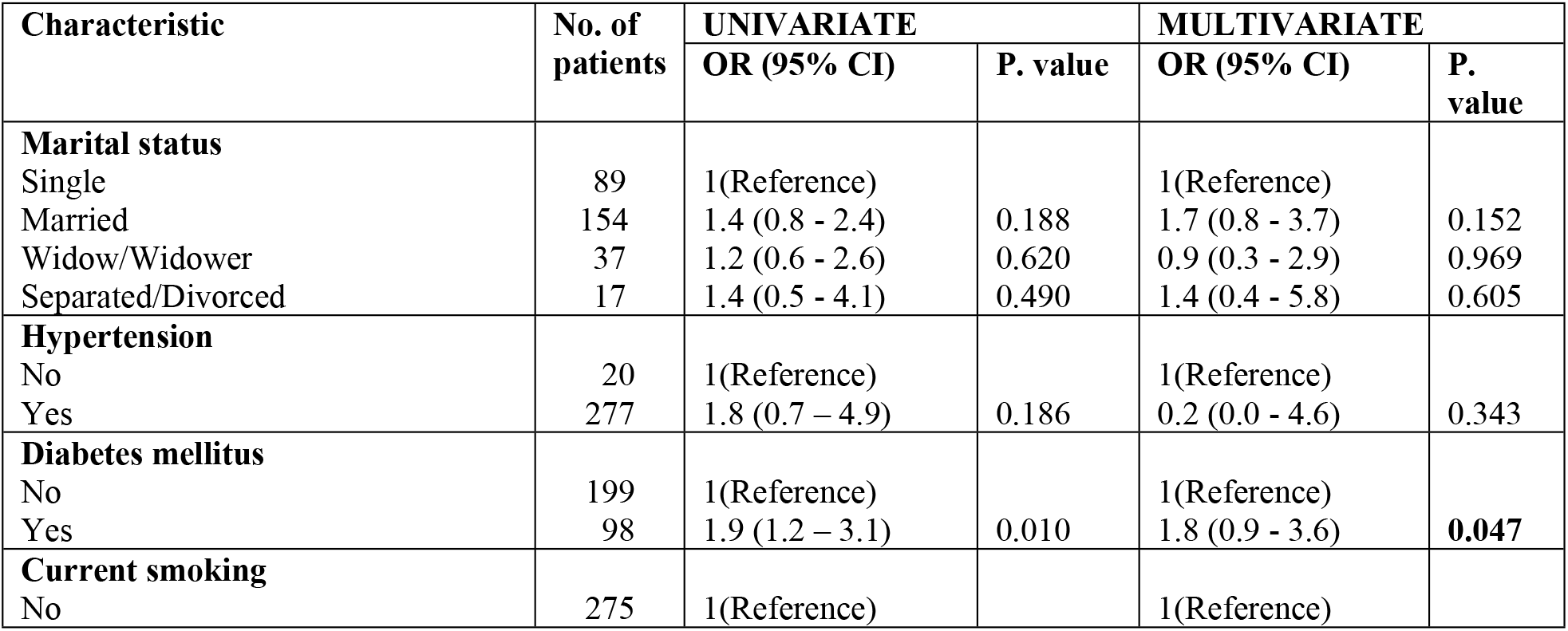
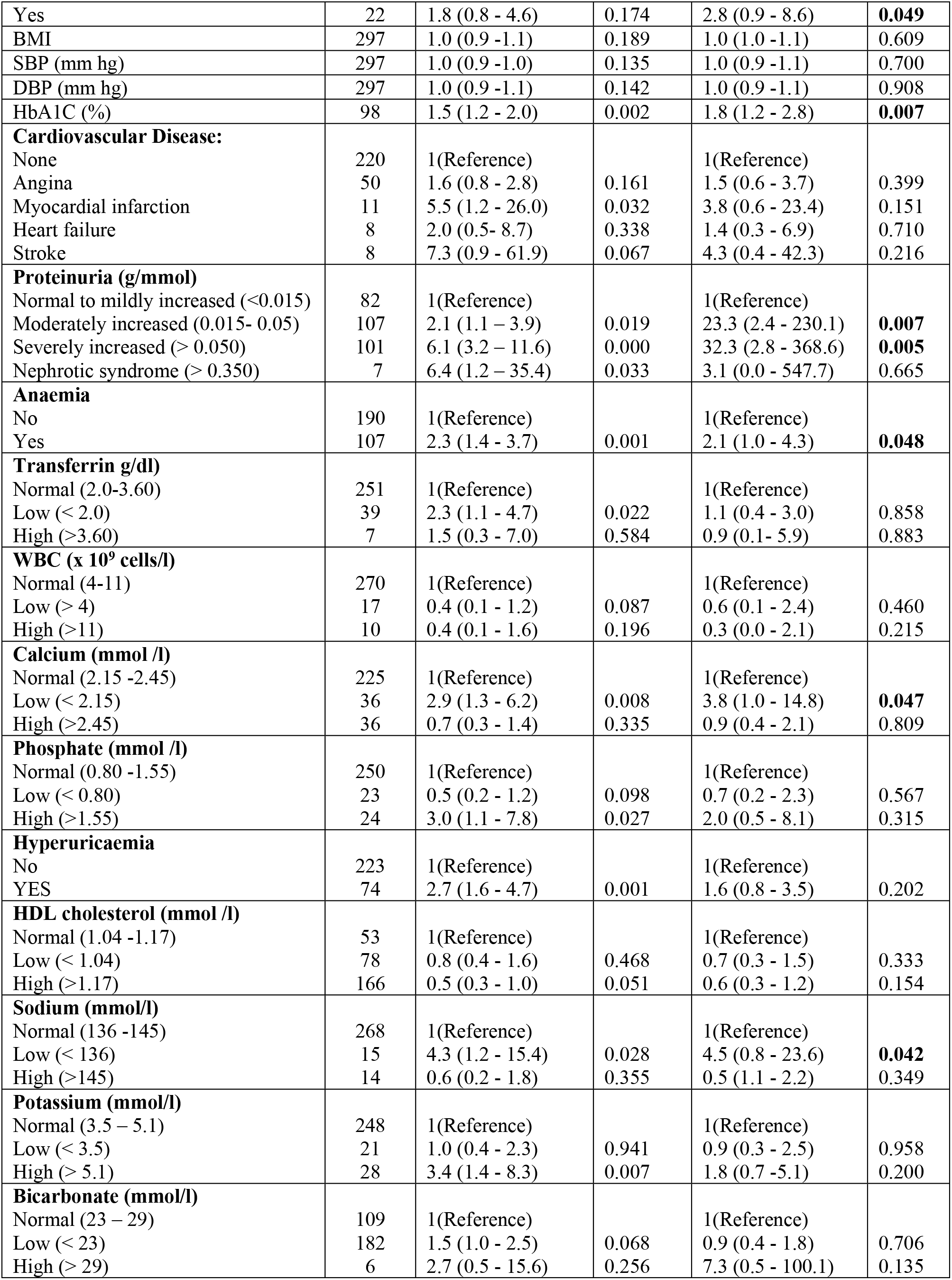
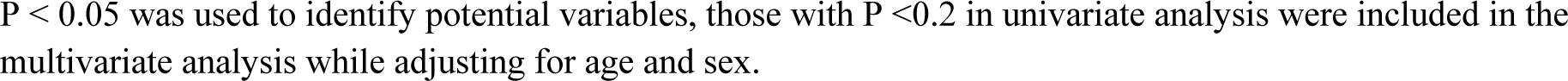
Predictors of CKD progression.

## Discussion

Our study identified a higher prevalence of progression of CKD in a prospective longitudinal study of black patients with CKD attending the kidney outpatient clinic at CMJAH in Johannesburg compared to literature reports. We found a high prevalence of CKD progression with 49.5% of the study patients with CKD progression, which is a higher prevalence of progression than the reported prevalence of 24% in a UK cohort study and 38 % in a study conducted in northern California in USA (27, 31). The possible explanation could be differences in the study populations; in our study we enrolled only black patients; black patients tend to have more rapid CKD progression even in the early stages of CKD (32, 33). Also, black patients due to racial/ethnic disparities are at higher risk of CKD progression and its related complications than other races (34, 35). This study established that 33% patients had CKD remission and 17.5% patients had CKD regression, similar to findings from other studies from the UK (31, 36). A sustained decline in eGFR of > 4 ml/min/1.73 m^2^/year or more was able to identify 48.5% patients who had CKD progression, including those with rapid CKD progression, similar to other studies (27, 37).

Hypertension was prevalent in the study cohort, occurring in 96.7 % of the study cohort at baseline and in 95.2% of CKD progressors and in 91.3% of those without progression. Of the patients with CKD progression, the majority (95.2%) had hypertension, 40.1% had diabetes mellitus and 39.5% had both hypertension and diabetes mellitus. Studies have reported CKD progression to be associated with CKD etiology and hypertension (38, 39). Both systolic and diastolic blood pressure were not significantly associated with CKD progression in this study, unlike findings from other studies where higher SBP and DBP levels were associated with CKD and had higher risks of CKD progression (40, 41). This difference could be due to the high prevalence (above 90%) of hypertension in both groups and an entry criterion of controlled blood pressure. Smoking was reported in 9.5 % of our study patients who had CKD progression, with a 2.8-fold increased risk for CKD progression; studies have reported that smoking is significantly associated with a higher risk of CKD and smoking is strongly associated with CKD progression (42, 43). Obesity and HDL cholesterol were not significantly associated with CKD progression in this study unlike findings from other studies where patients who had obesity and/or hyperlipidaemia had higher rates of CKD progression (44, 45). This difference could be due to the high prevalence of obesity/overweight in both progressor and non-progressor groups.

Diabetes mellitus had a 1.8-fold increased risk for CKD progression and increased HbA1c levels had a 1.8-fold increased risk for CKD progression similar to findings from other studies where increased HbA1c levels was associated with CKD progression in patients with T2DM (46, 47). Most (48.3%) patients with CKD progression presented with severely increased proteinuria; CKD progression was 32.3 times higher if a patient had severely increased proteinuria and 23.3 times higher if a patient had moderately increased proteinuria; studies have reported CKD progression to be increased in patients who present with higher levels of proteinuria (48, 49). ACEIs or ARBs have shown to reduce proteinuria more effectively than other antihypertensives; in this study, small numbers (20.2%) of patients were using ACEIs or ARBs, similar to other studies reporting underutilization of ACEIs/ARBs in CKD patients (50, 51).

Anaemia was reported in 45.6 % of patients who had CKD progression and was 2.1-fold increased with CKD progression and CKD was more severe if a patient had anaemia. Studies have reported that anaemia in CKD mainly results from impaired production of erythropoietin by the failing kidneys and anaemia is significantly associated with increased odds for CKD, CKD progression and worse clinical outcomes (52, 53). Hyperuricaemia was significantly associated with CKD progression on univariate analysis in our study but this significance was lost on multivariate analysis; studies have reported that high serum uric acid levels commonly occur as a result of the impaired glomerular filtration rate (GFR) that occurs in CKD. Hyperuricaemia can also precede the development of CKD, predict incident CKD and is associated with high risk for advanced CKD/ CKD progression (54, 55). Hypocalcaemia was reported in 17.7 % patients who had CKD progression with 3.8-fold association with CKD progression; low serum calcium levels are commonly caused by increased serum phosphorus and decreased renal production of 1,25(OH)2 vitamin D due to hyperparathyroidism. Hypocalcaemia is independently associated with CKD, and it tends to predict rapid CKD progression and severe CKD (56, 57). Hyperphosphataemia was significantly associated with CKD progression on univariate analysis but this significance was lost on multivariate analysis; studies have reported that high serum phosphate levels predicted higher rates of CKD progression (58, 59). Hyponatraemia was reported in 8.16 % of those patients who had CKD progression; CKD progression was 4.5 times more common if a patient had low serum sodium levels. The kidneys have the ability to modulate sodium and water excretion, and CKD is frequently complicated with hyponatraemia probably due to fluid overload or diuretic usage. Hyponatraemia in CKD is associated with a higher risk of CKD progression, cardiovascular events, hospitalization and increased mortality (60, 61). Hyperkalaemia was significantly associated with CKD progression on univariate analysis but this significance was lost on multivariate analysis. Studies have reported that high serum potassium levels are common in patients with chronic CKD and it is associated with decreased renal function and CKD progression; its prevalence increases as the eGFR declines (62, 63). Metabolic acidosis was not significantly associated with CKD progression in this study; studies from other settings reported that patients with low serum bicarbonate levels had higher rates of CKD progression and an accelerated reduction in eGFR with poor outcomes (64, 65). This difference could be due to the high prevalence of metabolic acidosis in both groups.

## Conclusion

Our study identified a higher prevalence of progression of CKD in a prospective longitudinal study of black patients with CKD compared with literature reports. Progression of CKD was associated with proteinuria, diabetes mellitus, elevated HbA1c, anaemia, hypocalcaemia, hyponatraemia and current smoking in a cohort of CKD patients with hypertension and diabetes mellitus. This is a call for nephrologists and clinicians to be vigilant in identifying CKD patients at risk of CKD progression at early stages as this would allow risk stratification to improve kidney disease outcomes. A sustained decline in eGFR of >4 ml/min/1.73 m^2^/year identified the highest number of patients with CKD progression, and this may help nephrologists and clinicians to identify those progressors in addition to capturing those patients with rapid CKD progression who require more frequent monitoring, closer surveillance and management towards halting CKD progression.

## Data Availability

Data cannot be shared publicly because of ethics policy at University of Witwatersrand, the participants signed a consent form, which states that data is exclusively available for professional research staff. Data are available to qualifying organizations and/or individuals from the chairperson of the Human Research Ethics Committee (Medical) of the University of the Witwatersrand, Johannesburg (‘Committee’) who is Dr. Clement Penny, who may be contacted by e-mail on) for researchers who meet the relevant ethics criteria for access to these data.

## Abbreviations

CKD: chronic kidney diseases;
CMJAH: Charlotte Maxeke Johannesburg Academic Hospital;
CKD-EPI: chronic kidney disease epidemiology collaboration;
ESKD: end stage kidney disease;
T2DM: Type 2 Diabetes Mellitus;
eGFR: estimated glomerular filtration rate;
IDMS: isotope dilution mass spectroscopy;
uPCR: urine protein creatinine ratio;
CCBs: calcium channel blockers;
ACEIs: angiotensin converting enzyme inhibitors;
ARBs: Aldosterone receptors blockers;
KDIGO: kidney disease improving global outcomes;
NCDs: non-communicable diseases;
SSA: sub Saharan Africa;
UK: United Kingdom;
USA: United States of America

## Acknowledgements

Special thanks to all patients attending the Charlotte Maxeke Johannesburg Academic Hospital (CMJAH) KOPD clinic for their willingness to participate in this study and to the staff at the CMJAH KOPD clinic and laboratory for their continued care, proper keeping of patient data and support which made this study possible. I thank the International Society of Nephrology (ISN), the University of the Witwatersrand, the University of Dodoma and Shinei industries (Aichi, Japan) Co. Ltd, for support, and providing a good and conducive environment for my training.

## Authors’ contributions

Design of the work: AJM, SN, RD & GP.

Data collection: AJM, SN, RD & GP.

Data processing & analysis: AJM, SN, RD & DB.

Manuscript writing: AJM, SN, RD, GP, & DB

All authors have read and approved the final manuscript.

